# Risk scores and coronary artery disease in patients with suspected acute coronary syndrome and intermediate cardiac troponin concentrations

**DOI:** 10.1101/2024.04.30.24306662

**Authors:** Daniel Perez-Vicencio, Alexander JF Thurston, Dimitrios Doudesis, Rachel O’Brien, Amy V. Ferry, Takeshi Fujisawa, Michelle C. Williams, Alasdair J. Gray, Nicholas L Mills, Kuan Ken Lee

**Affiliations:** BHF/University Centre for Cardiovascular Science, University of Edinburgh, Edinburgh, UK; Usher Institute, University of Edinburgh, Edinburgh, UK; Edinburgh Heart Centre, Royal Infirmary of Edinburgh, Edinburgh, UK; Department of Emergency Medicine, Emergency Medicine Research Group, Royal Infirmary of Edinburgh, Edinburgh, UK; Edinburgh Imaging Facility, University of Edinburgh, Edinburgh, UK

**Author notes:** Contributed equally (Daniel Perez-Vicencio, Alexander JF Thurston). Corresponding Author: Dr Kuan Ken Lee BHF/University Centre for Cardiovascular Science The University of Edinburgh Edinburgh EH16 4SA United Kingdom.

**Keywords:** acute coronary syndrome, cardiac troponin, risk scores, CT coronary angiography, coronary artery disease

## Abstract

**Background:** Guidelines recommend the use of risk scores to select patients for further investigation after myocardial infarction has been ruled out but their utility to identify those with coronary artery disease is uncertain.

**Methods:** In a prospective cohort study, patients with intermediate high-sensitivity cardiac troponin I concentrations (5 ng/L to sex-specific 99^th^ percentile) in whom myocardial infarction was ruled out were enrolled and underwent coronary computed tomography angiography (CCTA) after hospital discharge. HEART, EDACS, GRACE, TIMI, SCORE2 and PCE risk scores were calculated and the odds ratio (OR) and diagnostic performance for obstructive coronary artery disease determined using established thresholds.

**Results:** In 167 patients enrolled (64±12 years, 28% female), 29.9% (50/167) had obstructive coronary artery disease. The odds of having obstructive disease was increased for all scores with the lowest and highest increase observed for an EDACS score ≥16 (OR 2.2 [1.1-4.6]) and a TIMI risk score ≥1 (OR 12.9 [3.0-56.0]), respectively. The positive predictive value (PPV) was low for all scores but was highest for a GRACE score >88 identifying 39% as high-risk for a PPV of 41.9% (30.4-54.2%). The negative predictive value (NPV) varied from 77.3% to 95.2% but was highest for a TIMI score of 0 identifying 26% as low-risk for a NPV of 95.2% (87.2-100%).

**Conclusions:** In patients with intermediate cardiac troponin concentrations in whom myocardial infarction has been ruled out, clinical risk scores can help identify patients with and without coronary artery disease, but the performance of established risk thresholds requires optimisation for this purpose.

**Clinical Trial Registration:** https://clinicaltrials.gov/study/NCT04549805

## Introduction

Current strategies to assess patients with suspected acute coronary syndrome in the Emergency Department primarily utilise high-sensitivity cardiac troponin testing and the electrocardiogram to rule in and rule out myocardial infarction [1, 2]. Single measurement rule-out pathways are optimised to identify patients with low cardiac troponin concentrations who can be safely discharged from the Emergency Department without further testing and those with elevated concentrations who require admission for further assessment [3–8]. However, 1 in 4 patients have intermediate cardiac troponin concentrations and whilst they can be ruled out or ruled in with a second troponin measurement, they remain at higher risk of future adverse cardiac events [5, 9–11]. Similarly serial testing pathways with cardiac troponin measurement at 0/1- and 0/2-hours stratify patients into three groups including an observe zone group with intermediate concentrations [12, 13]. Guidelines recommend further investigation is considered in these patients but the optimal approach to select these patients for investigation is unknown [2, 14].

Clinical risk scores are widely used in the Emergency Department to risk stratify patients with suspected acute coronary syndrome [15–18]. However, the utility of these risk scores is uncertain, particularly following the widespread adoption of high-sensitivity cardiac troponin as a risk stratification tool [19]. Derived from historical cohorts, risk scores may lack external validity when applied to contemporary practice [20], and some include elements of the history or clinician gestalt, that may be subjective or vulnerable to bias. Nevertheless, recent guidelines continue to recommend the use of clinical risk scores to select patients with intermediate cardiac troponin concentrations for further testing [1, 2].

In a secondary analysis of the PRECISE-CTCA study [21], we evaluate the performance of clinical risk scores to identify coronary artery disease in patients with intermediate cardiac troponin concentrations in whom myocardial infarction has been ruled out.

## Methods

### Study design and population

PRECISE-CTCA (Troponin to Risk Stratify Patients with Acute Chest Pain for Computed Tomography Coronary Angiography) was a prospective cohort study conducted at the Royal Infirmary of Edinburgh, United Kingdom, between December 4, 2018, and October 6, 2020, that has previously reported (ClinicalTrials.gov, number NCT04549805) [21]. Patients >30 years old presenting with suspected acute coronary syndrome, in whom myocardial infarction had been ruled out and peak cardiac troponin concentrations were within the normal reference range, were recruited in the Emergency Department [21]. Patients who were unable to undergo CCTA due to severe renal failure (estimated glomerular filtration rate <30 mL/min/1.73 m^2^) or major allergy to iodinated contrast media, clear alternative diagnosis, requirement for in-patient investigation, CCTA or invasive coronary angiogram within the past 1 year, pregnancy or breast feeding and inability to give informed consent were excluded. The South East Scotland Research Ethics Committee 01 approved this study, and all participants provided written informed consent.

Only patients with intermediate cardiac troponin concentrations (between 5 ng/L and the sex-specific 99^th^ percentile) were included in this secondary analysis. Cardiac troponin was measured using the ARCHITECT_STAT_ high-sensitivity cardiac troponin I assay (Abbott Laboratories, Abbott Park, Illinois). This assay has a limit of detection of 1.2 ng/L and an inter-assay co-efficient of variation of <10% at 4.7 ng/L, with a sex-specific upper reference limit or 99^th^ percentile of 16 ng/L in females and 34 ng/L in males [22]. According to current national and international recommendations, symptoms of angina were classified as typical, atypical, or nonanginal chest pain using the Diamond and Forrester questions [23].

### Coronary Computed Tomography Angiography

All participants underwent CCTA as an outpatient as soon as possible after their initial hospital attendance. CCTA was performed using a 128-detector row scanner (Biograph mCT, Siemens Healthcare, Germany) with iodine-based contrast media, as per Society of Cardiovascular Computed Tomography (SCCT) guidelines [24]. All CCTA images were analysed by trained observers who performed a per-segment analysis using a 15-segment model to assess coronary artery stenoses. Luminal cross-sectional area stenoses were classified as normal (<10%), mild non-obstructive (10%-49%), moderate non-obstructive (50%-70%), or obstructive (>70% in the ≥1 major epicardial artery or >50% in the left main stem). Patients were classified according to the most significant stenosis identified on CCTA irrespective of whether the vessel has been stented. Coronary stenoses that were bypassed by a vascular graft were not considered in the classification.

### Clinical risk scores

We calculated the HEART, EDACS, GRACE 2.0, TIMI, PCE, and SCORE2 risk scores in all patients and used established thresholds for each to stratify patients as low or high risk ***(Supplemental Text 1 and Supplemental Figure 1)*** [17, 25–31]. The HEART score assesses risk of major adverse cardiac events at 6 weeks in patients presenting with chest pain to the Emergency Department using a threshold of <3 to identify those who are low-risk [25]. The EDACS score assesses risk of major adverse cardiac events at 30 days in patients presenting with chest pain to the Emergency Department using a threshold of <16 to identify those who are low-risk [17]. The GRACE 2.0 score assesses risk of death or recurrent myocardial infarction at 6 months in patients with acute coronary syndromes identifying patients with a score >88 at increased risk [17]. TIMI score assess the risk of death, re-infarction or ischemic events at 14 days in patients with acute coronary syndrome with as score of ≥1 associated with increased risk [27]. The PCE predicts risk of a first atherosclerotic cardiovascular disease event at 10 years with the score associated with a <5% selected as low risk [28, 29]. SCORE2 and SCORE2-OP estimates the risk of fatal cardiovascular disease at 10 years in adults aged 40-69 and 70 years or older, respectively. The SCORE2 and SCORE2-OP risk scores are used together in this analysis and were calibrated for low-risk regions with the score associated with a <5% selected as low risk [30, 31] ***(Supplemental Text 1)***.

### Statistical analysis

Baseline characteristics were presented as mean ± standard deviation or median [interquartile range] for continuous variables and as count (%) for categorical variables. The Welch two-sample *t*-test test and one-way ANOVA were used to compare continuous variables while Fisher’s exact test was used to compare categorical variables. Multiple imputation by chained equation or Markov chain Monte Carlo method was performed to account for missing variables [32]. We multiple-imputed all missing values in the variables required to calculate clinical risk scores except for cardiac troponin concentrations ***(Supplemental Figure 2*)**. We evaluated the association between the clinical risk scores and presence of any coronary artery disease and obstructive coronary artery disease separately using binomial logistic regression modelling by obtaining the exponential of the logistic regression coefficient. We calculated the diagnostic performance for each clinical risk score with 95% confidence intervals of the sensitivity, specificity, negative predictive value, and positive predictive value based on the rule-in/rule-out thresholds. Overall diagnostic accuracy was evaluated by receiver operating characteristic (ROC) curve and area under the curve analyses. All calculations were performed for any coronary artery disease and obstructive disease separately. We subsequently performed a sensitivity analysis restricted to patients who were not previously known to have coronary artery disease ***(Supplemental Table 1)***. All data analysis were conducted in R (version 4.3.0, R Foundation for Statistical Computing).

## Results

### Study population

In this secondary analysis 167 patients (64±12 years, 28% female) were included with intermediate cardiac troponin I concentrations and a median maximal concentration of 8 ng/L (inter-quartile range 6-12 ng/L). Of these patients, 120 (72%) had coronary artery disease and 50 (30%) had obstructive coronary artery disease on CCTA **(*Table 1*).**

**Table 1.**
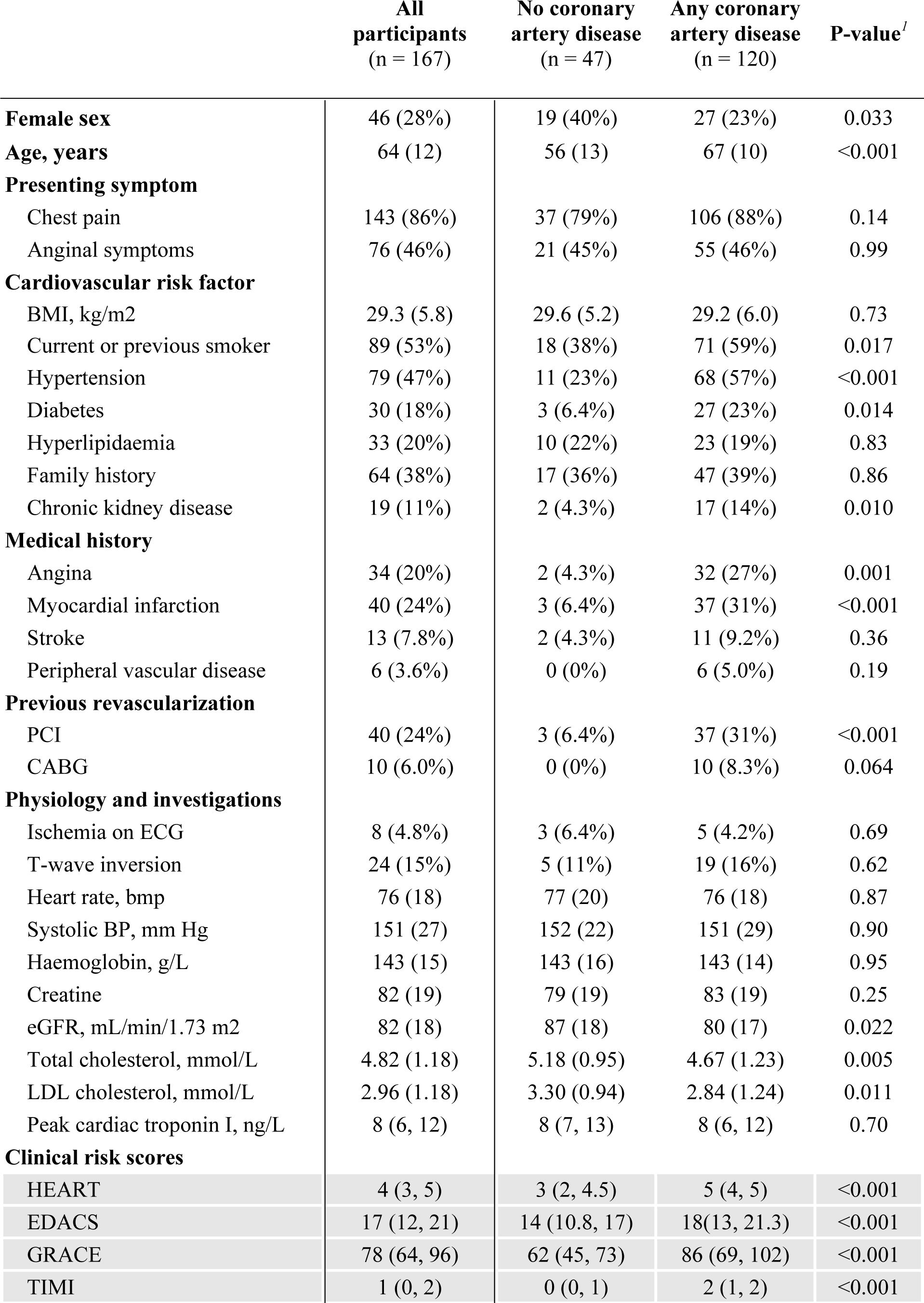
Baseline characteristics of patients with intermediate cardiac troponin concentrations stratified by findings on coronary computed tomography angiography.

Patients with coronary artery disease were older than those without (67±10 years *versus* 56±13 years, respectively; P<0.001) and more likely to be current or previous smokers (59% [71/120] *versus* 38% [18/47]; P=0.017). Patients with coronary artery disease were also more likely to have hypertension (57% [68/120] *versus* 23% [11/47]; P=0.017), diabetes mellitus (23% [27/120] *versus* 6.4% (3/47]; P=0.014) and chronic kidney disease (14% [17/120] *versus* 4.3% [2/47]; P=0.010) compared to those without **(*Table 1*)**. Similarly, patients with coronary artery disease were also more likely to have symptoms of typical angina (27% [32/120] *versus* 4.3% [2/47]; P=0.001) and to have both previous myocardial infarction (31% [37/120] *versus* 6.4% [3/47]; P<0.001) and percutaneous coronary intervention (31% [37/120] *versus* 6.4% [3/47]; P<0.001) than those without **(*Table 1*)**. Similar findings were observed when stratified according to the presence or absence of obstructive coronary artery disease ***(Supplemental Table 2)*.**

**Table 1.** Baseline characteristics of patients with intermediate cardiac troponin concentrations stratified by findings on coronary computed tomography angiography.

|  | **All**<br>**participants**<br>**(n = 167)** | **No coronary**<br>**artery disease**<br>**(n = 47)** | **Any coronary**<br>**artery disease**<br>**(n = 120)** | **P-value<sup>1</sup>** |
| --- | --- | --- | --- | --- |
| PCE | 0.19<br>(0.08, 0.36) | 0.09<br>(0.04, 0.16) | 0.26<br>(0.13, 0.38) | <0.001 |
| SCORE2/OP | 10.5 (5, 16) | 5 (4, 9) | 12.5 (7, 18) | <0.001 |

Values are median (interquartile range), n (%) or mean ± SD. <sup>1</sup> Pearson's Chi-squared test; Wilcoxon rank sum test; Fisher's exact test. BMI = body mass index; BP = blood pressure; bmp = beats per minute; CABG = coronary artery bypass grafting; CAD = coronary artery disease; ECG = electrocardiogram; EDACS = Emergency Department Assessment of Chest Pain Score; eGFR = estimated glomerular filtration rate; GRACE 2.0 = Global Registry of Acute Coronary Events version 2.0; HEART = History, ECG, Age, Risk factors, Troponin; LDL = low-density lipoprotein; PCE = pooled cohort equations; PCI = percutaneous coronary intervention; SCORE2/OP = Systematic COronary Risk Evaluation 2/Older Population; TIMI = Thrombolysis In Myocardial Infarction.

### Distribution of clinical risk scores

The median scores for patients with obstructive disease were significantly higher than those without coronary artery disease for all risk scores, HEART (5 ([inter-quartile range 4-5] *versus* 3 [2-4.5]; P<0.001), EDACS (18 [14–24] *versus* 14 [10.8-17]; P=0.002), GRACE (88 [77–108] *versus* 62 [45–73]; P<0.001), TIMI (2 [1–3] *versus* 0 [0–1]; P<0.001), PCE (0.32 [0.16-0.45] *versus* 0.09 [0.04-9]; P<0.001), and SCORE2 (15 ([8.5-18.8] *versus* 5 [4–9]; P<0.001), respectively **(*Figure 1*** ***& Supplemental Table 2)***. Similarly, patients with obstructive coronary artery disease had higher median scores than those with non-obstructive disease ***(Supplemental Table 2)*.**

**Figure 1.**
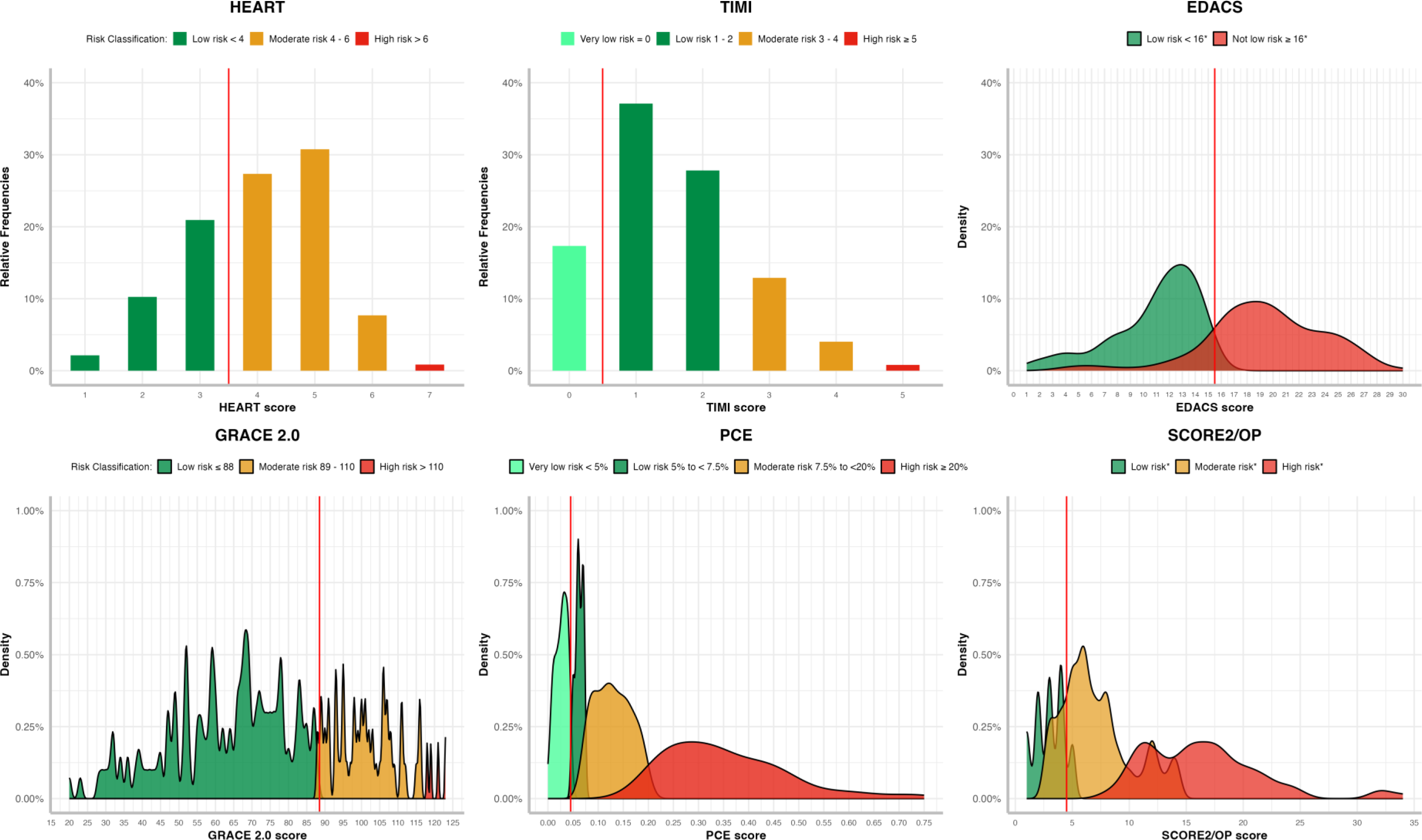
Distribution of risk scores in patients with suspected acute coronary syndrome and intermediate cardiac troponin concentrations stratified using established risk score thresholds as low-, moderate-, or high-risk. * When EDACS and SCORE2/OP are applied further criteria are recommended (Supplement Text 1) (8).

### Diagnostic performance of clinical risk scores

Patients with risk scores above the established risk threshold were more likely to have coronary artery disease than those below the risk threshold **(*Figure 2*)**. The odds ratio of having any coronary artery disease or obstructive disease was increased for all scores comparing those with increased scores to those with scores below the risk threshold. The odds ratio for obstructive coronary artery disease varied with the lowest increase observed for an EDACS score ≥16 (OR 2.2 [1.1-4.6]) and the highest increase for a TIMI risk score ≥1 (OR 12.9 [3.0-56.0]). Similarly, the odds ratio for any coronary artery disease varied with the lowest increase observed for an EDACS score ≥16 (OR 2.7 [1.3-5.3]) and the highest increase for a TIMI risk score ≥1 (OR 8.8 [4.0-19.2]), respectively **(*Figure 3*)**.

**Figure 2.**
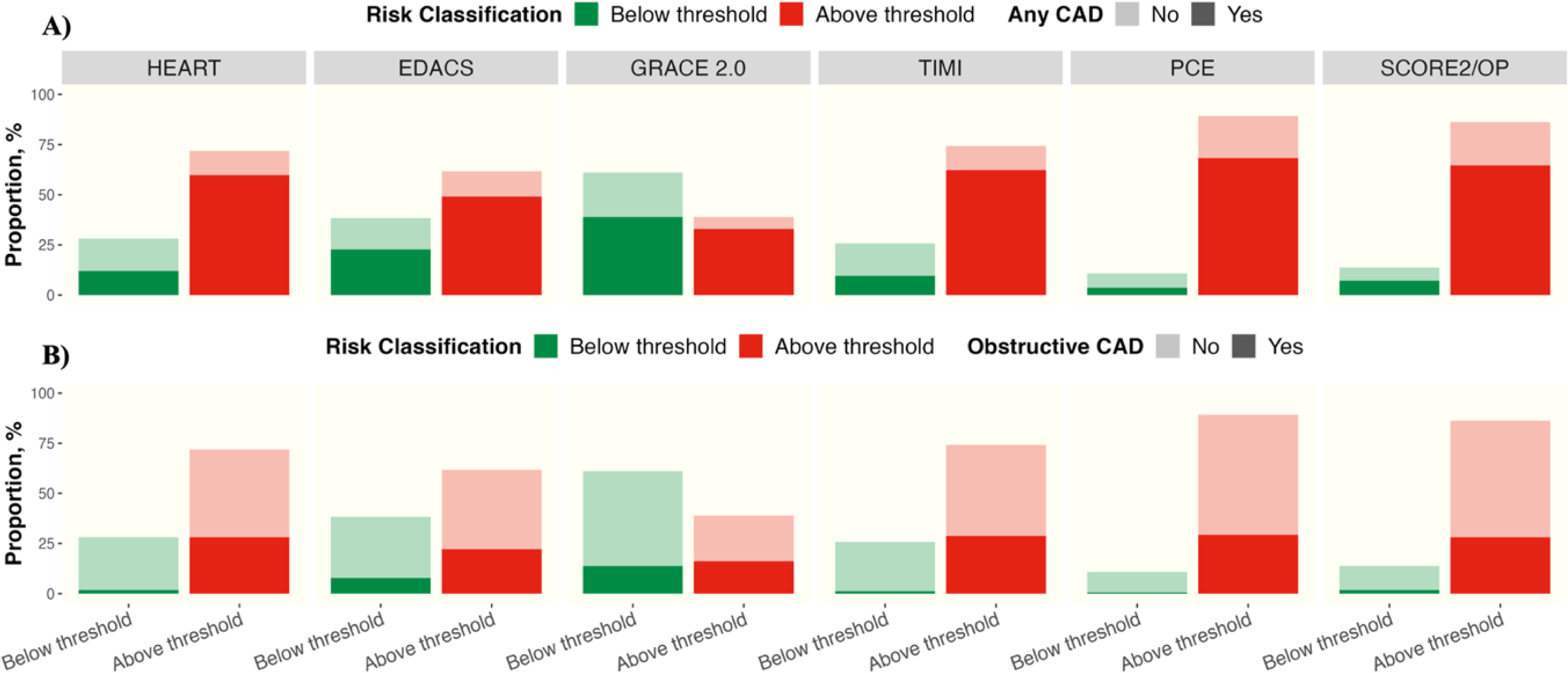
Proportion of patients with suspected acute coronary syndrome and intermediate cardiac troponin concentrations found to have any coronary artery disease (panel a) or obstructive disease (panel b) on CCTA below or above established low-risk thresholds for each risk score. * When EDACS and SCORE2/OP are applied further criteria are recommended (Supplemental Text 1). CAD = coronary artery disease, CCTA = coronary computer tomography angiography.

**Figure 3.**
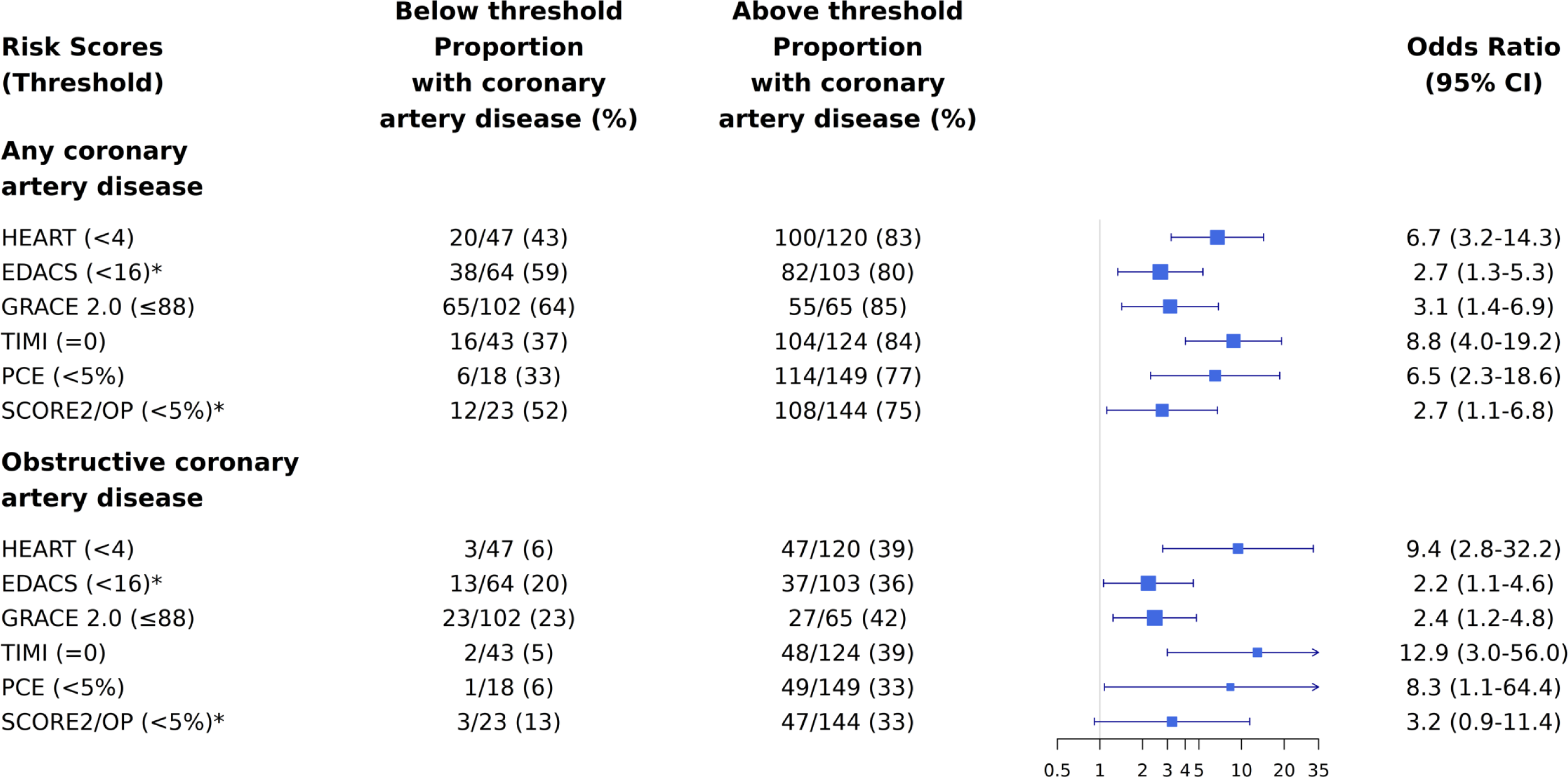
Odds ratio of having any or obstructive coronary artery disease on CCTA in patients with suspected acute coronary syndrome and intermediate cardiac troponin concentrations stratified by risk scores. Odds ratios for any coronary artery disease and obstructive disease on CCTA in patients with intermediate cardiac troponin concentrations comparing those with scores above and below established risk thresholds.

The positive predictive value (PPV) was low for all scores and the negative predictive value varied widely. Across all clinical risk scores, a GRACE 2.0 score of >88 had the highest positive predictive value for obstructive coronary artery disease identifying 39% as high-risk with a PPV of 41.9% (30.4-54.2% confidence interval). The negative predictive value (NPV) varied from 77.3% to 95.2% but was highest for a TIMI score of 0 identifying 26% as low risk with a NPV of 95.2% (87.2-100%) **(*Figure 4*** ***& Supplemental Table 3)***. Similar findings were observed when considering any coronary artery disease (***Supplementary Table 4)*.**

**Figure 4.**
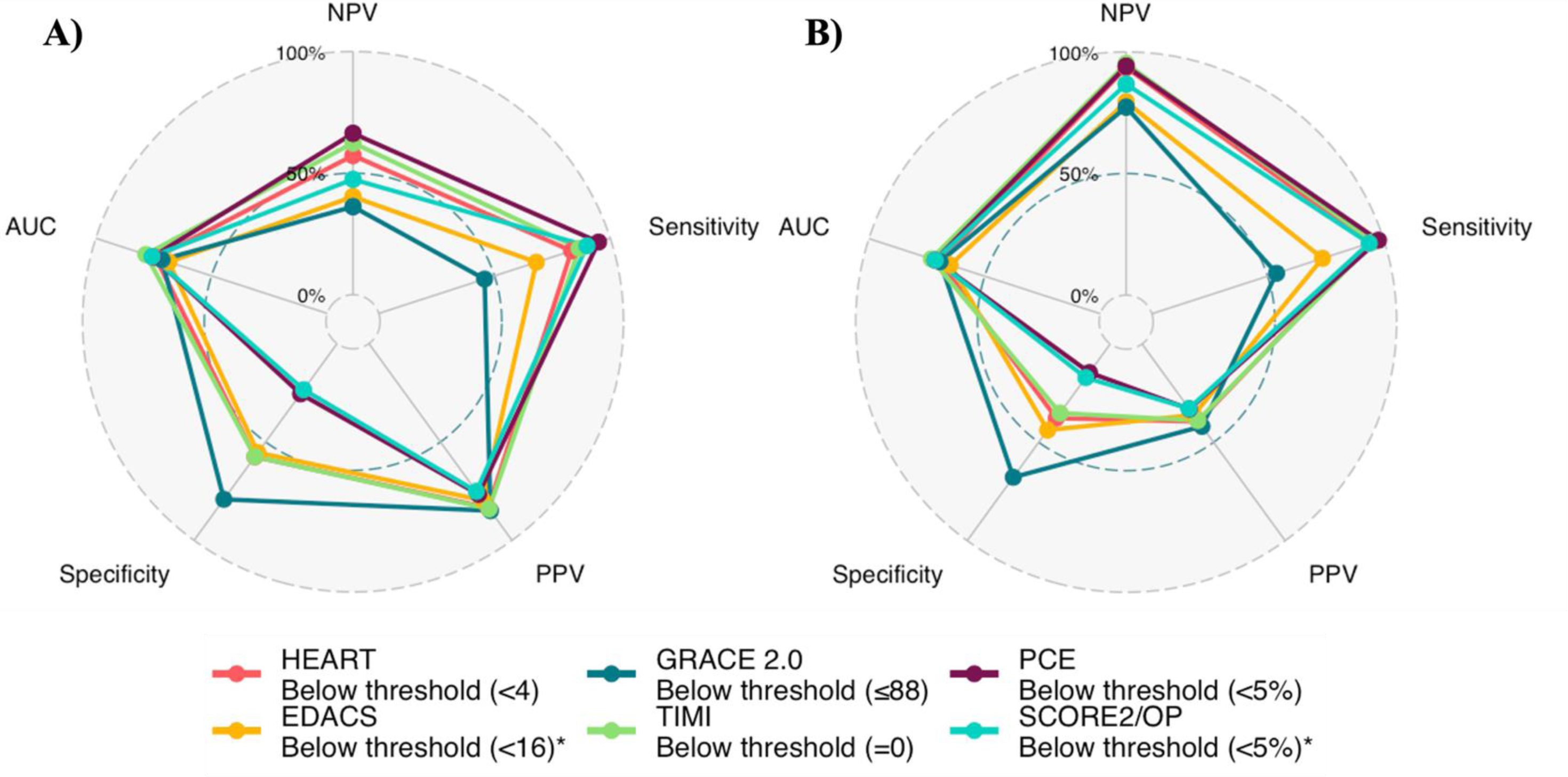
Radar plot comparing diagnostic performance of risk scores for any coronary artery disease (panel a) and obstructive disease (panel b) in patients with suspected acute coronary syndrome and intermediate cardiac troponin concentrations. AUC = area under the curve, NPV = negative predictive value, PPV = positive predictive value.

### High sensitivity troponin I *versus* clinical risk scores

All clinical risk scores had a higher discriminatory performance than high-sensitivity cardiac troponin alone (area under receiver operator curve [AUC] 0.481 [0.383-0.580 confidence interval] and 0.533 [0.440 – 0.625] for any coronary artery disease and obstructive disease, respectively). The TIMI risk score had the highest discrimination for coronary artery disease and obstructive disease (0.784 [0.713 - 0.854] and 0.730 [0.653 - 0.808], respectively). The lowest performing clinical risk score was EDACS for both coronary artery disease and obstructive disease (0.684 [0.597 - 0.772] and 0.649 [0.555 - 0.743], respectively***) (Figure 5*)**.

**Figure 5.**
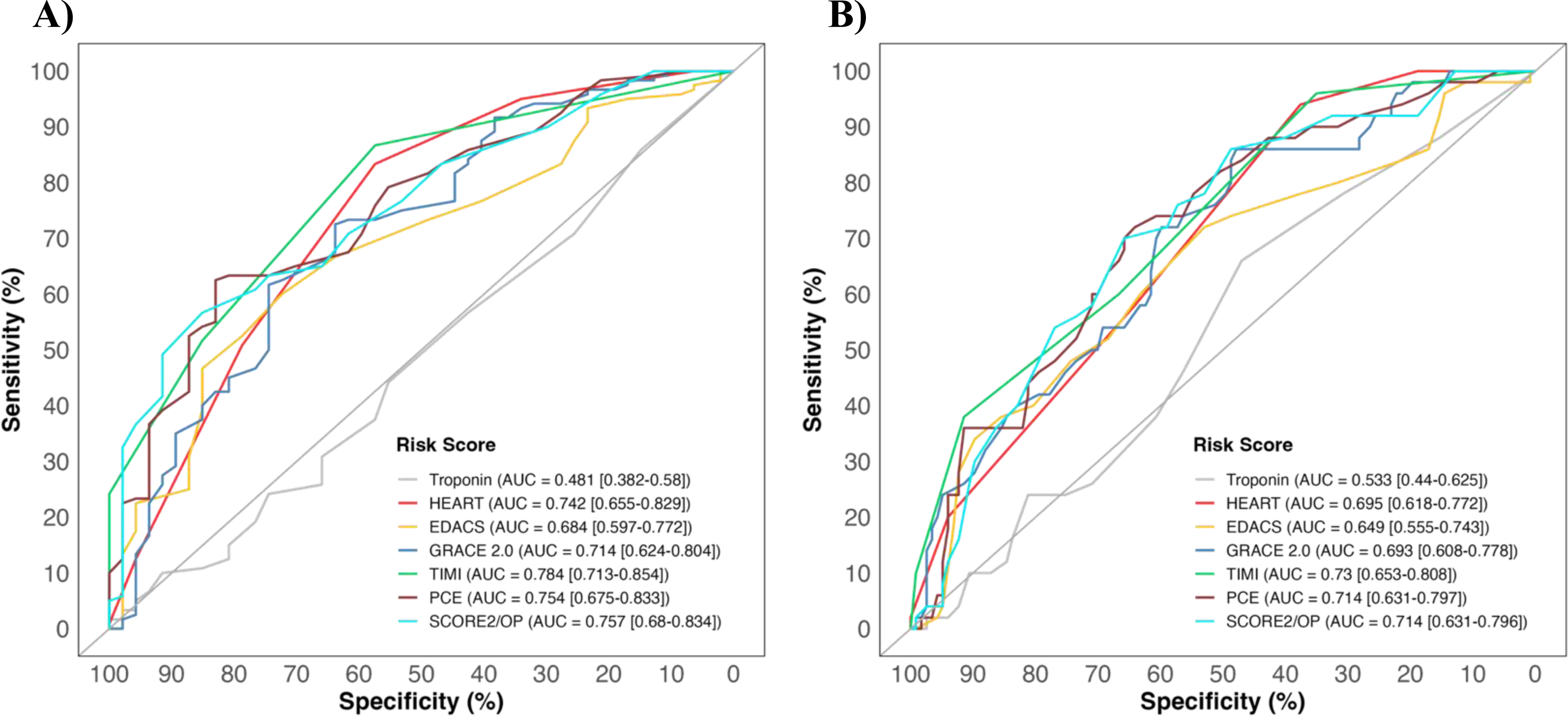
Discrimination of cardiac troponin and clinical risk scores for any coronary artery disease (panel a) and obstructive disease (panel b) in patients with suspected acute coronary syndrome and intermediate cardiac troponin concentrations. Receiver-operating-characteristic curves illustrating discrimination of cardiac troponin and clinical risk scores for coronary artery disease (panel a) and obstructive disease (panel b) in patients with suspected acute coronary syndrome and intermediate cardiac troponin concentrations.

### Patients without previous diagnosis of coronary artery disease

In a sensitivity analysis restricted to the 103 (62%) patients not previously known to have coronary artery disease, the discrimination for coronary artery disease and obstructive disease of all risk scores was higher than cardiac troponin alone (cardiac troponin, AUC = 0.472 [0.383–0.580] and 0.584 [0.440–0.625], respectively). Discrimination was greatest for SCORE2 for the outcome of any coronary artery disease (0.753 [0.659 - 0.846] and for the PCE for the outcome of obstructive disease (0.747 [0.639 - 0.856]). EDACS again had the lowest discrimination for coronary artery disease and obstructive disease (0.658 [0.551-0.764] and 0.641 [0.488-0.795], respectively) ***(Supplemental Figure 3 and Supplemental Tables 5 & 6)*.**

## Discussion

In this study, we evaluated the performance of six established clinical risk scores to determine whether they could help to identify patients with intermediate troponin levels who are more likely to have coronary artery disease after myocardial infarction has been ruled out in the Emergency Department. We found that all risk scores improve the odds of identifying patients with coronary artery disease on CCTA. Using the existing risk threshold for each score, the positive predictive value is low for all, with the best performing being the GRACE 2.0 score which correctly identified 4 in 10 patients with obstructive disease. The negative predictive value was also low, with the best performing score being TIMI which correctly identifies 19 of 20 patients as not having obstructive disease.

Patients with suspected acute coronary syndrome are at risk of future myocardial infarction or cardiac death even after myocardial infarction has been ruled out therefore clinical guidelines recommend further non-invasive investigations to identify potential underlying coronary artery disease [5, 14]. CCTA has been suggested as the non-invasive investigation modality of choice due to its ability to accurately assess coronary artery plaque burden and characteristics to guide use of secondary preventative therapies such as antiplatelets and statins to modify their risk of future major adverse cardiovascular outcomes [2, 33–35]. However, given resource constraints and the large volume of patients presenting with suspected acute coronary syndrome, it would be valuable to develop strategies to select patients with a higher pre-test probability of coronary artery disease to avoid unnecessary CCTA. In our previous analysis, we demonstrated that high-sensitivity cardiac troponin I can help identify patients with a higher prevalence of coronary artery disease for further testing after myocardial infarction has been ruled out. Those with intermediate cardiac troponin concentrations had 3-times higher odds of having coronary artery disease compared to those with very low troponin concentrations. However, this remains a substantial group of patients, comprising approximately 1 in 4 of all patients with suspected acute coronary syndrome in whom cardiac troponin testing alone does not further discriminate those who are likely to have coronary artery disease. Strategies to refine risk in this group of patients could therefore help target further investigations more judiciously.

Multiple risk scores have been developed and validated for the initial triage of patients with suspected acute coronary syndrome and for the risk stratification of apparently healthy individuals [5, 17, 21, 25–31]. These risk scores are recommended by clinical guidelines to guide early referral for specialist investigation such as invasive coronary angiography or initiation of preventative medications [2, 14]. Given that these risk scores incorporate known cardiovascular risk factors and were primarily developed to predict risk of major adverse cardiovascular outcomes, it is perhaps not surprising that they also improve the identification of patients with coronary artery disease in the Emergency Department. Nevertheless, none of the risk scores we have evaluated had optimal rule-in and rule-out performance for any or obstructive coronary artery disease and implementing multiple risk scores for this purpose would be challenging in practice. Developing novel risk stratification tools specifically for this group of patients could overcome this current limitation of existing risk scores.

We acknowledge there are limitations to our analysis. We did not have high-sensitivity cardiac troponin T measurements for this cohort and so could not include an analysis of the Troponin-only Manchester Acute Coronary Syndromes score [36]. The EDACS and TIMI scores use a previous history of varying degrees of coronary artery disease as components of the score, which may inflate their performance. However, in a sensitivity analysis restricted to patients without previously known coronary artery disease, the negative predictive value and positive predictive value for these scores remained similar. While the risk scores evaluated here were designed to predict risk of short- or long-term clinical outcomes rather than to diagnose coronary artery disease, this is the underlying pathophysiological basis of the majority of adverse cardiovascular events, and the diagnosis of coronary artery disease is important to patients with chest pain and can facilitate the targeting of preventative therapies that could reduce risk of these outcomes.

## Conclusions

In patients with intermediate cardiac troponin concentrations in whom myocardial infarction has been ruled out, clinical risk scores can help identify patients with and without obstructive coronary artery disease, but the performance of established risk thresholds requires optimisation for this purpose.

## Data Availability

The R code and anonymised data used for analysis can be made available to researchers on request to the corresponding author

https://github.com/dvicencio/RiskScorescvd

## Non-standard Abbreviations and Acronyms

CCTA: Coronary Computed Tomography Angiography
HEART: History, Electrocardiogram, Age, Risk factors, and Troponin
EDACS: Emergency Department Assessment of Chest Pain Score
TIMI: Thrombolysis in Myocardial Infarction
GRACE: 2.0 Global Registry of Acute Coronary Events version 2.0
ASCVD-PCE: Atherosclerosis Cardiovascular Disease – Pooled Cohort Equations
SCORE2: Systematic COronary Risk Evaluation 2
MACE: Major Adverse Cardiac Events

## Acknowledgements

The authors gratefully acknowledge the BHF Cardiovascular Biomarker Laboratory, University of Edinburgh, for their assistance with this work.

## References

1. Collet, J.P., et al., 2020 ESC Guidelines for the management of acute coronary syndromes in patients presenting without persistent ST-segment elevation. Eur Heart J, 2021. 42(14): p. 1289–1367.

2. Writing Committee, M., et al., 2021 AHA/ACC/ASE/CHEST/SAEM/SCCT/SCMR Guideline for the Evaluation and Diagnosis of Chest Pain: A Report of the American College of Cardiology/American Heart Association Joint Committee on Clinical Practice Guidelines. J Am Coll Cardiol, 2021. 78(22): p. e187–e285.

3. Shah, A.S.V., et al., High-sensitivity troponin in the evaluation of patients with suspected acute coronary syndrome: a stepped-wedge, cluster-randomised controlled trial. Lancet, 2018. 392(10151): p. 919–928.

4. Body, R., et al., Rapid exclusion of acute myocardial infarction in patients with undetectable troponin using a high-sensitivity assay. J Am Coll Cardiol, 2011. 58(13): p. 1332–9.

5. Shah, A.S., et al., High-sensitivity cardiac troponin I at presentation in patients with suspected acute coronary syndrome: a cohort study. Lancet, 2015. 386(10012): p. 2481–8.

6. Chapman, A.R., et al., Comparison of the Efficacy and Safety of Early Rule-Out Pathways for Acute Myocardial Infarction. Circulation, 2017. 135(17): p. 1586–1596.

7. Anand, A., et al., High-Sensitivity Cardiac Troponin on Presentation to Rule Out Myocardial Infarction: A Stepped-Wedge Cluster Randomized Controlled Trial. Circulation, 2021. 143(23): p. 2214–2224.

8. Jaffe, A.S., et al., Single Troponin Measurement to Rule Out Myocardial Infarction: JACC Review Topic of the Week. J Am Coll Cardiol, 2023. 82(1): p. 60–69.

9. Allen, B.R., et al., Diagnostic Performance of High-Sensitivity Cardiac Troponin T Strategies and Clinical Variables in a Multisite US Cohort. Circulation, 2021. 143(17): p. 1659–1672.

10. Chapman, A.R., et al., Association of High-Sensitivity Cardiac Troponin I Concentration With Cardiac Outcomes in Patients With Suspected Acute Coronary Syndrome. JAMA, 2017. 318(19): p. 1913–1924.

11. Bularga, A., et al., High-Sensitivity Troponin and the Application of Risk Stratification Thresholds in Patients With Suspected Acute Coronary Syndrome. Circulation, 2019. 140(19): p. 1557–1568.

12. Cullen, L., et al., Validation of high-sensitivity troponin I in a 2-hour diagnostic strategy to assess 30-day outcomes in emergency department patients with possible acute coronary syndrome. J Am Coll Cardiol, 2013. 62(14): p. 1242–1249.

13. Reichlin, T., et al., One-hour rule-out and rule-in of acute myocardial infarction using high-sensitivity cardiac troponin T. Arch Intern Med, 2012. 172(16): p. 1211–8.

14. Byrne, R.A., et al., 2023 ESC Guidelines for the management of acute coronary syndromes. Eur Heart J Acute Cardiovasc Care, 2023.

15. Mahler, S.A., et al., The HEART Pathway randomized trial: identifying emergency department patients with acute chest pain for early discharge. Circ Cardiovasc Qual Outcomes, 2015. 8(2): p. 195–203.

16. Than, M.P., et al., Effectiveness of EDACS Versus ADAPT Accelerated Diagnostic Pathways for Chest Pain: A Pragmatic Randomized Controlled Trial Embedded Within Practice. Annals of Emergency Medicine, 2016. 68(1): p. 93–102.

17. Than, M., et al., Development and validation of the Emergency Department Assessment of Chest pain Score and 2 h accelerated diagnostic protocol. Emergency Medicine Australasia, 2014. 26(1): p. 34–44.

18. Flaws, D., et al., External validation of the emergency department assessment of chest pain score accelerated diagnostic pathway (EDACS-ADP). Emergency Medicine Journal, 2016. 33(9): p. 618–625.

19. Chapman, A.R., et al., High-Sensitivity Cardiac Troponin I and Clinical Risk Scores in Patients With Suspected Acute Coronary Syndrome. Circulation, 2018. 138(16): p. 1654–1665.

20. Balasubramanian, R.N., et al., Role and relevance of risk stratification models in the modern-day management of non-ST elevation acute coronary syndromes. Heart, 2023. 109(7): p. 504-+.

21. Lee, K.K., et al., Troponin-Guided Coronary Computed Tomographic Angiography After Exclusion of Myocardial Infarction. J Am Coll Cardiol, 2021. 78(14): p. 1407–1417.

22. Shah, A.S., et al., High sensitivity cardiac troponin and the under-diagnosis of myocardial infarction in women: prospective cohort study. BMJ, 2015. 350: p. g7873.

23. Diamond, G.A. and J.S. Forrester, Analysis of Probability as an Aid in the Clinical-Diagnosis of Coronary-Artery Disease. New England Journal of Medicine, 1979. 300(24): p. 1350–1358.

24. Abbara, S., et al., SCCT guidelines for the performance and acquisition of coronary computed tomographic angiography: A report of the society of Cardiovascular Computed Tomography Guidelines Committee Endorsed by the North American Society for Cardiovascular Imaging (NASCI). Journal of Cardiovascular Computed Tomography, 2016. 10(6): p. 435–449.

25. Mahler, S.A., et al., The HEART Pathway Randomized Trial: Identifying Emergency Department Patients with Acute Chest Pain for Early Discharge. Circulation, 2014. 130.

26. Elbarouni, B., et al., Validation of the Global Registry of Acute Coronary Event (GRACE) risk score for in-hospital mortality in patients with acute coronary syndrome in Canada. American Heart Journal, 2009. 158(3): p. 392–399.

27. Scirica, B.M., et al., Validation of the thrombolysis in myocardial infarction (TIMI) risk score for unstable angina pectoris and non-ST-elevation myocardial infarction in the TIMI III registry. Am J Cardiol, 2002. 90(3): p. 303–5.

28. Goff, D.C., Jr., et al., 2013 ACC/AHA guideline on the assessment of cardiovascular risk: a report of the American College of Cardiology/American Heart Association Task Force on Practice Guidelines. J Am Coll Cardiol, 2014. 63(25 Pt B): p. 2935–2959.

29. Rana, J.S., et al., Accuracy of the Atherosclerotic Cardiovascular Risk Equation in a Large Contemporary, Multiethnic Population. J Am Coll Cardiol, 2016. 67(18): p. 2118–2130.

30. Hageman, S., et al., SCORE2 risk prediction algorithms: new models to estimate 10-year risk of cardiovascular disease in Europe. European Heart Journal, 2021. 42(25): p. 2439–2454.

31. de Vries, T.I., et al., SCORE2-OP risk prediction algorithms: estimating incident cardiovascular event risk in older persons in four geographical risk regions. European Heart Journal, 2021. 42(25): p. 2455–2467.

32. Jakobsen, J.C., et al., When and how should multiple imputation be used for handling missing data in randomised clinical trials - a practical guide with flowcharts. Bmc Medical Research Methodology, 2017. 17.

33. Williams, M.C., et al., Coronary Artery Plaque Characteristics Associated With Adverse Outcomes in the SCOT-HEART Study. J Am Coll Cardiol, 2019. 73(3): p. 291–301.

34. Andelius, L., et al., Impact of statin therapy on coronary plaque burden and composition assessed by coronary computed tomographic angiography: a systematic review and meta-analysis. Eur Heart J Cardiovasc Imaging, 2018. 19(8): p. 850–858.

35. Haase, R., et al., Diagnosis of obstructive coronary artery disease using computed tomography angiography in patients with stable chest pain depending on clinical probability and in clinically important subgroups: meta-analysis of individual patient data. BMJ, 2019. 365: p. l1945.

36. Body, R., et al., Troponin-only Manchester Acute Coronary Syndromes (T-MACS) decision aid: single biomarker re-derivation and external validation in three cohorts. Emerg Med J, 2017. 34(6): p. 349–356.

